# Unveiling the spectrum of Respiratory Syncytial Virus disease in Adults: from Community to Hospital

**DOI:** 10.1101/2024.10.17.24315581

**Authors:** Koos Korsten, Matthijs R.A. Welkers, Thijs van de Laar, Alex Wagemakers, Peter van Hengel, Peter C. Wever, Eva Kolwijck

**Affiliations:** Department of Medical Microbiology and Infection Control, Amsterdam UMC, location AMC, Amsterdam, Netherlands; Department of Medical Microbiology Onze Lieve Vrouwe Gasthuis, Amsterdam, The Netherlands; Department of Pulmonary Diseases, Flevo Hospital, Almere, the Netherlands; Department of Medical Microbiology and Infection Prevention, Jeroen Bosch Hospital, ‘s Hertogenbosch, the Netherlands

## Abstract

**Background:** Respiratory syncytial virus can cause severe disease in the older adult population. Three vaccines for RSV are currently market approved but the risk of RSV-hospitalization in (older) adults from a community level remains elusive. We aimed to estimate the risk of RSV-hospitalization and characterize the patients that end up in hospital.

**Methods:** We manually analyzed records of adults aged ≥20 with RSV-infection between 2022-2024 in three hospitals in the Netherlands. These hospitals implemented routine RSV-testing at emergency departments. Using population-based data in combination with the in-hospital data, we estimated the population risk of RSV-hospitalization. Hospital records were analyzed to characterize the role RSV played in their course of disease.

**Results:** We analyzed 709 RSV cases of whom 503 (70.9%) were hospitalized. 526 patients were ≥60, and 183 were <60 years of age. The population risk of RSV-hospitalization was 0.006-0.02% for patients aged 20-59 years and 0.04-0.24% for those ≥60. The highest risks were seen in older patients with congestive heart disease (0.14-5.0%) and COPD (0.17-1.76%). RSV caused clinically relevant infection in 88% of hospitalized cases but was only mentioned using specific ICD-codes in 4.4%. Comorbidity was prevalent (88.5%) and exacerbation of underlying disease caused of 46.3% of RSV-related hospital admissions. ICU admittance was 11.2% and in-hospital mortality was 8.1%.

**Conclusion:** The risk of RSV-hospitalization from the community is low but is increased substantially in those with underlying disease. RSV is often clinically relevant in hospitalized patients by causing exacerbation of underlying disease but is infrequently mentioned in specific ICD-codes.

## INTRODUCTION

Respiratory Syncytial Virus (RSV) is a leading cause of respiratory infections worldwide [1]. RSV has emerged as a substantial contributor to respiratory morbidity and mortality in the aging population [2]. RSV therapeutics are limited to ribavirin and RSV vaccine development has proved challenging as multiple attempts were unsuccessful and even harmful [1]. The discovery of the neutralization-specific viral epitopes of the pre-fusion RSV glycoprotein have boosted the search for effective RSV vaccines [3]. Three vaccines have recently shown to be effective [4-6] and have been granted U.S. Food and Drug Administration and European Medicines Agency approval [7-10]. However, it remains unclear who would truly benefit most from these vaccines. It is insufficiently known how many people in the general population are infected with RSV and what fraction is subsequently hospitalized. Furthermore, age and presence of underlying comorbidity are intertwined but it is important to know the role of each of these factors. There are several limitations in the current data that could impact estimation of the disease burden in (older) adults [11]. Serious outcomes such as the hospitalization rate in the general population are rare requiring large studies. Even the large vaccine trials could not provide this estimated as they were limited by low RSV-incidence rates [4-6]. Another major factor resulting in potential underestimation of the burden of RSV-related disease is the use of strict testing definitions based on clinical symptoms e.g. fever, that may not be sensitive for RSV [12]. The lack of routine testing in clinical practice is a major limitation that affects studies based on health claims databases, as diagnostic codes for RSV are only present if testing was performed [11]. In patients with comorbidities, RSV might be the trigger for hospitalization but may not be the registered hospital diagnosis which could also underestimate the role of RSV in these hospitalizations. Last, younger adults are not considered at risk for severe RSV disease, but individuals with underlying comorbidity might still develop serious sequela that require hospitalization.

RSV vaccines are entering the market while the indications and cost-effectiveness still needs to be determined. With this study, we aim to provide insight in the proportion of patients in the community that require hospitalization due to RSV. Characteristics of these patients are determined to evaluate what drives subsequent hospitalization and clinical course to determine who might benefit from RSV vaccination.

## METHODS

### Patients and testing procedures

RSV testing results were retrospectively collected from adults aged ≥20 years that visited the ED of the three participating hospitals in the Netherlands between January 2022 and April 2024. We chose this time period since the epidemiology of RSV was disturbed during the actual COVID-19 outbreak. We included adults ≥20 years to align with population-based age data. The included hospitals were the Amsterdam University Medical Center (tertiary teaching hospital, Amsterdam), Jeroen Bosch Hospital (large general hospital, ‘s Hertogenbosch) and the Flevo Hospital (general hospital, Almere). All patients that tested positive for RSV were included. Patients with nosocomial infection, defined as a positive test at least seven days after hospital admission were excluded. To estimate the proportion of patients who had acquired RSV in the community and required hospitalization, we selected two of the participating hospitals based on their defined ‘catchment area’. This included the Jeroen Bosch Hospital which serves the area of ‘s Hertogenbosch, and the Flevo Hospital which serves the area of Almere. Both hospitals are situated in isolated geographical areas without other hospitals nearby which ensures that patients from those ‘catchment areas’ who needed inpatient care were most likely referred to those hospitals. All hospitals had implemented routine testing for SARS-CoV-2 since the start of the COVID-19 pandemic in 2020 using the GeneXpert CoV-2/Flu/RSV test [Cepheid, Sunnyvale, CA]. This resulted in ongoing RSV testing in all patients presenting with respiratory symptoms.

### Data collection

Population-based demographic data for the predefined catchment areas were collected using the Dutch census data from the StatLine database of Statistics Netherlands (CBS, “Centraal Bureau voor de Statistiek”) [13]. Statistics Netherlands is the principal statistical agency in the Netherlands. We collected regional data on age distribution [14] and national data from 2022-2023 on the prevalence of Chronic Obstructive Pulmonary Disease (COPD), asthma, diabetes and congestive heart disease (CHD) [15]. The estimates of the prevalence of comorbidities were extracted from the Netherlands Institute for Health Services Research (NIVEL) data, which is a sentinel surveillance system based on a comprehensive network of approximately 140 general practitioner practices across the Netherlands [15].

To determine the characteristics and assess clinical severity of patients that presented in hospital with RSV infections we manually analyzed the electronic patient file (EPD) data. We determined general patient characteristics including age, sex and comorbidities. We scored the presence of the following comorbidities; pulmonary disease (asthma/COPD/interstitial lung disease; ILD), CHD, metabolic disease (diabetes, renal insufficiency/dialysis), hematologic and solid tumor malignancy, rheumatologic/immunologic/auto-immune disease and organ transplantation with active immunosuppression therapy. Secondly, we checked the ED status to extract the reason of presentation, vital signs, infection markers (C-reactive protein (CRP) and leukocyte count) and radiographic evaluation. In patients admitted to hospital, we subsequently determined the reason for hospitalization, length of stay, culture results, prescription of antibiotics, intensive care unit (ICU) admittance, need for supplementary oxygen or invasive respiratory support and mortality. In order to determine the presence of an bacterial superinfection we created a scoring system which involved the presence of radiologic signs of pneumonia (2 points), relevant positive microbiologic culture result (2 points), enhanced infection markers (CRP >100 mg/L and/or leukocyte count >11×10^9^/L, 1 point) and antibiotic therapy prescribed for presumed pulmonary bacterial superinfection (0,5 points). A bacterial superinfection score below two was classified as “unlikely”, scores ranging from 2-3 were classified as “possible” while a score >3 deemed a bacterial superinfection “likely”. We classified the role that RSV played in course of disease based on the EPD. We defined the categories ‘primary RSV infection’, ‘exacerbation of underlying disease caused by RSV’, ‘Bacterial superinfection following RSV’ and ‘RSV considered irrelevant’. The classification was scored by the authors based on the full hospital records, admission and discharge letters. Relevance of RSV was determined based on presence of respiratory symptoms and/or if RSV was mentioned by the physicians as a relevant factor in the course of clinical disease. In case of an unclear diagnosis supplemental Table S1 was used to determine the most likely role of RSV and disagreement was solved by consensus between the authors. Last, we collected ICD-10 codes for hospitalized patients to quantify potential underreporting.

### Statistical analysis

The estimation of RSV-hospitalization incidence was modelled using population-based data combined with the in-hospital data. Demographic data from the aforementioned registries were used to obtain the number of people per age group in the predefined study regions (Flevo Hospital Almere and Jeroen Bosch Hospital, ‘s Hertogenbosch). The estimates of the community-based RSV incidence previous studies were used (5,4-7,2% RSV incidence in older adults ≥60; Korsten et al.[16] and 7% for healthy adults aged 18-60 years; Hall et al. [17]). Combined with the number of people in the population we extrapolated the number of patients that would theoretically be infected with RSV in the community seasonally. With the in-hospital data of proven RSV-positive patients in this study, we subsequently estimated the proportion of patients from the community that required hospitalization for RSV infection. Seasons were defined to calculate the incidence of RSV-hospitalization per RSV season. The seasons were defined retrospectively based on a period of three months before and after the peak incidence month of RSV-hospitalization during that year. We performed the same modeling exercise stratified for patients with COPD, asthma, CHD and diabetes. Next, we characterized the clinical severity of RSV positive in-hospital patients and compared patients that visited the ED who were, or were not, subsequently admitted to hospital. We did not use imputation to correct for missing data. All analyses were performed in R software, version 4.0.1 (www.r-project.org).

Ethical approval for the study was obtained from the Medical Ethics Committee Brabant (reference number NW2024-23) and individual patient informed consent was waived.

## RESULTS

### Population risk of RSV-hospitalization

From January 2022 up to April 2024 we extracted 45651 individual RSV test results collected from 32056 inpatients aged ≥20 years. After exclusion of nosocomial cases (n=53), 708 individual patients with 709 RSV infections remained, of whom 526 (74%) were aged ≥60 years and 183 were between 20-59 years old [Table 1]. One sixty-year old patient experienced two RSV infections separated by a four month interval and was therefore included twice. The majority (99.4%) of positive samples were from nasopharyngeal swabs collected in the ED. Two RSV seasons were defined, the 2022-2023 and 2023-2024 season both spanning from September until April based on peak incidence months in December each year. The RSV epidemiology in our study is shown in Figure 1.

**Table 1.** Number of respiratory syncytial virus (RSV) cases among total number of individual patients tested.

| Site | RSV season | Age 20-59<br>(RSV/total) | Age 60+<br>(RSV/total) |
| --- | --- | --- | --- |
| Amsterdam University Medical Center | 2022-2023 | 43/2013 (2.1%) | 87/2864 (3.3%) |
|  | 2023-2024 | 44/1182 (3.7%) | 65/1683 (3.9%) |
|  | Off-season | 21/3915 (0.5%) | 36/4916 (0.7%) |
| Jeroen Bosch Hospital | 2022-2023 | 26/921 (2.8%) | 136/2891 (4.7%) |
|  | 2023-2024 | 13/566 (2.3%) | 65/1631 (4.0%) |
|  | Off-season | 5/922 (0.5%) | 46/2771 (1.7%) |
| Flevo Hospital | 2022-2023 | 15/582 (2.6%) | 57/1032 (5.5%) |
|  | 2023-2024 | 11/534 (2.1%) | 25/844 (3.0%) |
|  | Off-season | 5/1058 (0.5%) | 9/1731 (0.5%) |
| **Total** | 2022-2024 | 183/11693 (1.6%) | 526/20363 (2.6%) |

Off-season is the periods between 1<sup>st</sup> January 2022 and 1<sup>st</sup> of April 2024 excluding the two defined RSV seasons

**Figure 1.**
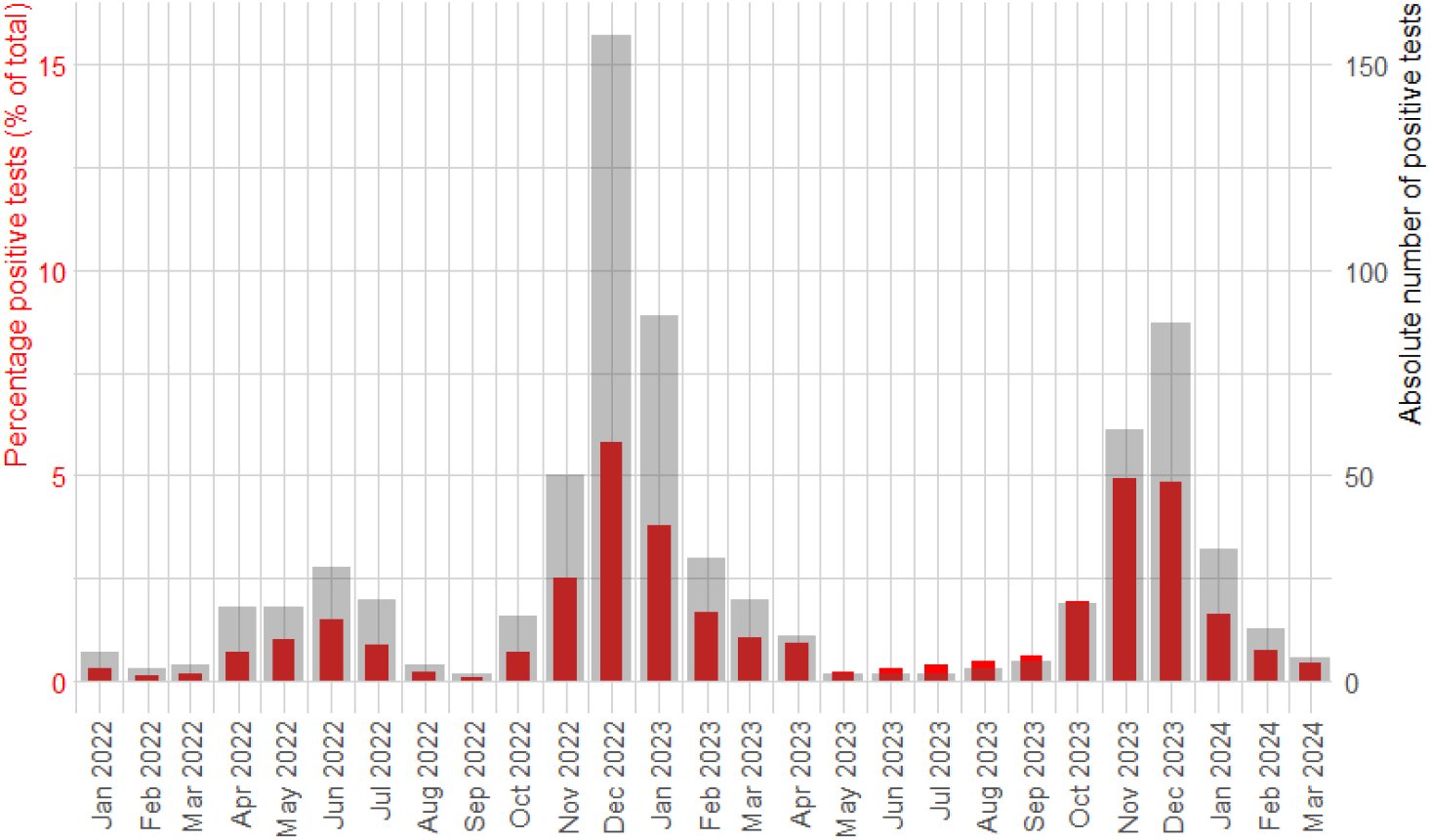
Epidemiology of respiratory syncytial virus (RSV) during the study period for all hospitals combined. The red bars define the percentage of positive RSV tests as a fraction of all performed RSV tests (scale on the left). The grey bars define the absolute number of positive RSV tests (scale on the right).

In total, 709 RSV episodes were recorded from the ED of whom 503 (70.9%) were subsequently admitted. This resulted in a population risk of RSV-hospitalization during the RSV season of 0.006-0.02% for patients aged 20-59 years [Table S2] and 0.04-0.24% for those aged ≥60 years [Table 2]. Based on the estimation of the theoretical number of RSV cases in the population, once positive for RSV, the risk of subsequent hospitalization was 0.08-0.29% in patients aged 20-59 years [Table S2] and 0.6-4.4% in those aged ≥60 [Table 2]. Subgroup analysis for comorbidities in those aged ≥60 is shown in Table 3. The highest population risks were seen in patients with CHD (0.14-5.0%) and COPD (0.17-1.76%). Estimates for patients aged 20-59 years with comorbidities are shown in supplemental Table S3. Older age increased the population risk of RSV-hospitalization with a maximal range of 0.002% in those aged 20-39 to 0.718% in those ≥90 [Figure 2, Table S4].

**Table 2.** Respiratory syncytial virus (RSV) epidemiology in adults aged 60 years and older.

|  |  | Region Flevo Hospital |  | Region Jeroen Bosch Hospital |  |
| --- | --- | --- | --- | --- | --- |
|  |  | Season 2022-2023 | Season 2023-2024 | Season 2022-2023 | Season 2023-2024 |
| **Population level** | Number of people in the population $\geq 60$<sup>1</sup> | 42166 | 44107 | 39807 | 40760 |
|  | Estimated RSV incidence (n. 5.4-7.2%)<sup>2</sup> | 2277-3036 | 2382-3176 | 2150-2866 | 2201-2935 |
| **Emergency department** | RSV cases (n) | 57 | 25 | 136 | 65 |
|  | Incidence from population<sup>3</sup> | 0.14% | 0.06% | 0.34% | 0.16% |
|  | Incidence from RSV positive (%)<sup>4</sup> | 1.9-2.5% | 0.8-1.0% | 4.7-6.3% | 2.2-3.0% |
| **Hospitalized** | RSV cases (n) | 47 | 19 | 94 | 57 |
|  | Incidence from population<sup>3</sup> | 0.11% | 0.04% | 0.24% | 0.14% |
|  | incidence from RSV positive (%)<sup>4</sup> | 1.5-2.1% | 0.6-0.8% | 3.3-4.4% | 1.9-2.6% |

<sup>1</sup> Based on regional data [14]. <sup>2</sup> Estimated based on RSV incidence in adults aged >60 years (10) <sup>3</sup> Number of confirmed RSV cases in this study visiting the ED/hospital divided by the total number of people in the population \*100%. <sup>4</sup> Number of confirmed RSV cases in this study visiting the ED/hospital divided by the number of estimated RSV cases in the population \*100%

**Table 3.** Respiratory syncytial virus (RSV) epidemiology in adults aged 60 years and older with comorbidity.

|  |  | Region Flevo hospital |  | Region Jeroen Bosch hospital |  |
| --- | --- | --- | --- | --- | --- |
|  |  | 2022-2023 | 2023-2024 | 2022-2023 | 2023-2024 |
| **Population level** | Number of people in the population $\geq 60$<sup>1</sup> | 42166 | 44107 | 39807 | 40760 |
|  | Patients with COPD (5.7-10.9%)<sup>2</sup> | 2403-4596 | 2514-4808 | 2269-4339 | 2323-4443 |
|  | Patients with asthma (9.4-10.2%)<sup>2</sup> | 3964-4301 | 4146-4499 | 3742-4060 | 3831-4158 |
|  | Patients with CHD (1.1-9.9%)<sup>2</sup> | 464-4174 | 485-4367 | 438-3941 | 448-4035 |
|  | Patients with diabetes (11.7-22.9%)<sup>2</sup> | 4933-9656 | 5161-10101 | 4657-9116 | 4769-9334 |
| **Emergency department RSV cases** | COPD: n(incidence<sup>3</sup>) | 15 (0.33-0.62%) | 8 (0.17-0.32%) | 52 (1.20-2.29%) | 24 (0.54-1.03%) |
|  | Asthma: n(incidence<sup>3</sup>) | 7 (0.16-0.17%) | 2 (0.04-0.05%) | 12 (0.30-0.32%) | 9 (0.22-0.23%) |
|  | CHD: n(incidence<sup>3</sup>) | 7 (0.17-1.51%) | 7 (0.16-1.44%) | 30 (0.76-6.85%) | 18 (0.41-4.0%) |
|  | Diabetes: n(incidence<sup>3</sup>) | 17 (0.18-0.34%) | 4 (0.04-0.08%) | 30 (0.33-0.64%) | 17 (0.18-0.36%) |
| **Hospitalized RSV cases** | COPD: n(incidence<sup>3</sup>) | 14 (0.30-0.58%) | 8 (0.17-0.32%) | 40 (0.92-1.76%) | 22 (0.50-0.95%) |
|  | Asthma: n(incidence<sup>3</sup>) | 6 (0.14-0.15%) | 2 (0.04-0.05%) | 6 (0.15-0.16%) | 8 (0.19-0.21%) |
|  | CHD: n(incidence<sup>3</sup>) | 6 (0.14-1.29%) | 6 (0.14-1.24%) | 22 (0.56-5.0%) | 17 (0.38-3.79%) |
|  | Diabetes: n(incidence<sup>3</sup>) | 13 (0.13-0.26%) | 3 (0.03-0.06%) | 18 (0.20-0.39%) | 17 (0.18-0.36%) |

COPD = chronic obstructive pulmonary disease. CHD = congestive heart disease. <sup>1</sup> Based on regional data [14]. <sup>2</sup> Prevalence based on the number of persons with that comorbidity per 1000 persons in the population. The lower and upper limits define the range in prevalence within specific age groups in those over 60 years of age [15]
<sup>3</sup> Number of confirmed RSV cases in this study visiting the ED/hospital with that comorbidity divided by the total number of people in this population \*100%

**Figure 2.**
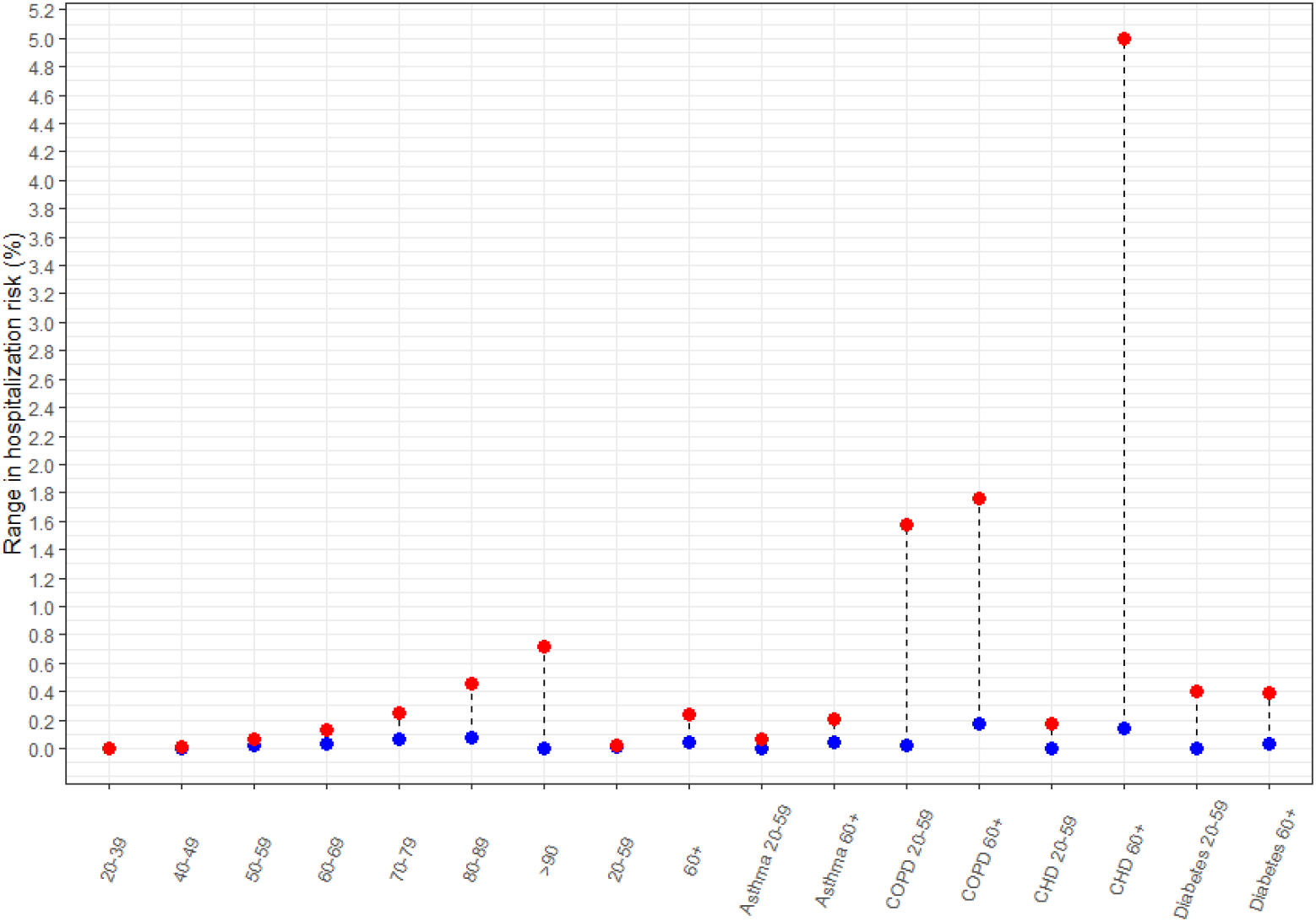
Hospitalization risk from the community setting for various age and comorbidity groups. The blue dot represent the lower estimate while the red dot represents the upper estimate.

### In-hospital severity

In total, 709 RSV episodes were examined based on the hospital records. Patient characteristics are shown in Table 4 and stratified per site in Table S5. Underlying comorbidity was present in 83.6%. The most frequent comorbidities were pulmonary (45.3%) and comorbidity was generally more prevalent in the older age group with the exception of rheumatic/immunologic disease which was more common in the younger age group. Cough was reported in the ED by 85% of patients while 74.7% of older adults and 55.7% of younger adults reported dyspnea. Fever was reported in 38%, and confirmed at the ED in only 29.9% of cases. Hospitalization was more frequent in older adults as compared to young adults (76.2% versus 55.7% respectively). There were differences in cohort characteristics per site [Table S5]. There was a higher percentage of cases from the tertiary teaching hospital with underlying malignancy and organ transplant and a higher fraction of patients that were <60 years as compared to the general hospitals. The main difference between the two general hospitals was that patients in the Jeroen Bosch hospital were relatively older (mean age 74 versus 68) and had more frequent CHD (23.0% versus 13.9%) as compared to those from the Flevo Hospital. This age difference aligns with the age distribution based on the demographic data of the areas [14].

**Table 4.** Characteristics of the study cohort.

|  | **Age 20-59**<br>**n=183** | **Age ≥60**<br>**n=526** | **Total**<br>**N=709** |
| --- | --- | --- | --- |
| Male | 82 (44.8%) | 249 (47.3%) | 331 (46.7%) |
| Age (median, [IQR]) | 47 [35-55] | 74 [67-82] | 69 [59-79] |
| Any comorbidity<sup>1</sup> | 139 (76.0%) | 454 (86.3%) | 593 (83.6%) |
| Pulmonary disease | 69 (37.7%) | 252 (47.9%) | 321 (45.3%) |
| - COPD | - 21 (11.5%) | - 178 (33.8%) | - 199 (28.1%) |
| - Asthma | - 36 (19.7%) | - 54 (10.3%) | - 90 (12.7%) |
| Congestive heart disease (CHD) | 15 ( 8.2%) | 126 (24.0%) | 141 (19.9%) |
| Cardiopulmonary disease<sup>2</sup> | 4 (2.2%) | 47 (8.9%) | 51 (7.2%) |
| Active malignancy | 31 (16.9%) | 95 (18.1%) | 127 (17.9%) |
| - Hematologic | - 17 ( 9.3%) | - 49 ( 9.5%) | - 66 ( 9.3%) |
| - Solid tumor | - 14 ( 7.7%) | - 47 ( 8.9%) | - 61 ( 8.6%) |
| Metabolic disease | 35 (19.1%) | 182 (34.6%) | 217 (30.5%) |
| - Diabetes | - 14 ( 7.7%) | - 130 (24.7%) | - 144 (20.3%) |
| - Chronic renal insufficiency | - 11 (6.0%) | - 54 (10.2%) | - 65 (9.2%) |
| - Hemodialysis | - 5 (2.7%) | - 10 (1.9%) | - 15 ( 2.1%) |
| Organ transplant<sup>3</sup> | 7 ( 3.8%) | 8 ( 1.5%) | 15 ( 2.1%) |
| Rheumatic / immunologic disease<sup>3</sup> | 37 (20.2%) | 62 (11.8%) | 99 (14.0%) |
| Symptoms (anamnestic) |  |  |  |
| - Cough | - 149 (81.4%) | - 454 (86.3%) | - 603 (85.0%) |
| - Dyspnea | - 102 (55.7%) | - 393 (74.7%) | - 495 (69.9%) |
| - Fever | - 83 (45.4%) | - 187 (35.6%) | - 270 (38.0%) |
| Fever measured in the ED (≥38°C) | 50 (29.6%) | 153 (29.9%) | 203 (29.9%) |
| CT value (median, [IQR]) | 27.5 [22-32] | 24.5 [21-29] | 24.8 [21-30] |
| Hospitalization | 102 (55.7%) | 401 (76.2%) | 503 (70.9%) |

<sup>1</sup>. From the comorbidities that were noted in the method section <sup>2</sup> Patients that both have CHD and pulmonary comorbidity <sup>3</sup>. With active immunosuppressive therapy or illness.

In 623/709 (88%) episodes, RSV was judged to be a clinically relevant finding in the course of the ED visit or subsequent hospital admission. Clinical severity in episodes in which RSV was clinically relevant is shown in Table 5. Hospitalized patients were older and had higher prevalence of comorbidity (88.5%) of which pulmonary disease was most prevalent (54.8%), followed by CHD (22.4%), diabetes (21.9%), and active malignancy (15.4%). Admittance to an ICU was needed in 11.2% of cases of whom 50% needed invasive respiratory support. In-hospital mortality was 8.1% in patients in whom RSV was clinically relevant. Stratified for age mortality was 9,3% in those above 60 years of age and 2,7% in younger adults [Table S6]. ICU admission was inversely more frequent in younger adults in which ICU admission was more often deemed feasible. RSV was most often associated with exacerbation of underlying disease in hospitalized patients (46.3%) while in patients that did not require hospital admission, RSV was often judged to be the sole cause of disease (68.8%). Treatment for presumed bacterial superinfection was given in 293 hospitalized cases of whom only 62 (21.2%) had a positive bacterial culture result [Table 5]. From the 116 cases suspected for bacterial superinfection following RSV, only 38.8% (45/116) had microbiological proof with a positive bacterial culture result. Based on the bacterial superinfection score 141/293 patients were likely to have an actual bacterial superinfection based on radiographic imaging, culture, inflammatory markers and prescribed antibiotics [Table S7].

**Table 5.** Severity of infection in those with clinically relevant respiratory syncytial virus infection (RSV)

|  | **Emergency department**<br>**n=189** | **Hospitalized**<br>**n=434** | **Deceased\***<br>**N=35** |
| --- | --- | --- | --- |
| Male | 91 (48.1%) | 192 (44.2%) | 17 (48.6%) |
| Age (median, [IQR]) | 64 [49-74] | 71.5 [63-81] | 79 [72-86] |
| - 20-39 | 23 (12.3%) | 15 (3.5%) | - 0 (0%) |
| - 40-59 | 48 (25.7%) | 61 (14.1%) | - 2 (5.7%) |
| - 60-79 | 93 (49.7%) | 233 (53.8%) | - 16 (45.7%) |
| - 80 and older | 23 (12.3%) | 124 (28.6%) | - 17 (48.6%) |
| Any comorbidity<sup>1</sup> | 152 (80.4%) | 384 (88.5%) | 30 (85.7%) |
| Pulmonary disease | 72 (38.1%) | 238 (54.8%) | 19 (54.3%) |
| - COPD | 35 (18.5%) | 160 (36.9%) | 16 (45.7%) |
| - Asthma | 29 (15.3%) | 58 (13.4%) | 2 (5.7%) |
| Congestive heart disease (CHD) | 28 (14.8%) | 97 (22.4%) | 12 (34.3%) |
| Cardiopulmonary disease<sup>2</sup> | 9 (4.8%) | 39 (9.0%) | 6 (17.1%) |
| Active malignancy | 46 (24.3%) | 67 (15.4%) | 5 (14.3%) |
| - Hematologic | - 24 (12.7%) | - 37 (8.5%) | - 3 (8.6%) |
| - Solid tumor | - 24 (12.7%) | - 28 (6.5%) | - 2 (5.7%) |
| Metabolic disease | 46 (24.3%) | 139 (32%) | 11 (31.4%) |
| - Diabetes | - 29 (15.3%) | - 95 (21.9%) | - 10 (28.6%) |
| - Chronic renal insufficiency | - 14 (7.4%) | - 38 (8.8%) | - 2 (5.7%) |
| - Hemodialysis | - 6 (3.2%) | - 7 (1.6%) | - 0 (0%) |
| Organ transplant<sup>3</sup> | 5 (2.6%) | 6 (1.4%) | 0 (0%) |
| Rheumatic / immunologic disease<sup>3</sup> | 31 (16.4%) | 57 (13.1%) | 2 (5.7%) |
| **Severity** |  |  |  |
| Length of stay for total hospital admission<br>median days [IQR] (all hospitalized patients) | NA | 5 [3-8] | 6 [2-10] |
| median days [IQR] (ICU admitted patients) | NA | 10 [8-23] | 6 [2-30] |
| Infiltrate found with chest radiography | NA | 164 (37.9%) | 17 (48.6%) |
| Bacterial culture positive | NA | 62 (21.2%) | 5 (19.2%) |
| Treated with antibiotics<sup>4</sup> | NA | 293 (68%) | 26 (74.3%) |
| Oxygen therapy n(%)<br>median days [IQR] | NA | 331 (76.6%)<br>3 [2-7] | 33 (94.3%)<br>3 [1-7] |
| Intensive care unit admission n(%)<br>median days in the ICU [IQR] | NA | 48 (11.2%)<br>4 [2-11] | 11 (31.4%)<br>6 [2-10] |
| Invasive ventilation n(%)<br>median days [IQR] | NA | 24 (5.6%)<br>6 [3-9] | 10 (28.6%)<br>6 [2-8] |
| Role of RSV |  |  |  |
| - Primary RSV infection | - 130 (68.8%) | - 112 (25.8%) | - 6 (17.1%) |
| - Factor in exacerbation of underlying disease | - 50 (26.5%) | - 201 (46.3%) | - 17 (48.6%) |
| - Bacterial superinfection after RSV | - 9 (4.8%) | - 116 (26.7%) | - 9 (25.7%) |
| - Indeterminate<sup>5</sup> | - 0 (0%) | - 5 (1.2%) | - 3 (8.6%) |

NA = not applicable, IQR = interquartile range \* Deceased patients are also included in the hospitalized group.
<sup>1</sup>. From the comorbidities that were noted in the method section <sup>2</sup>. Patients that both have CHD and pulmonary comorbidity <sup>3</sup> With active immunosuppressive therapy or illness. <sup>4</sup>. For presumed respiratory bacterial superinfection. <sup>5</sup>. No definitive role of RSV could be addressed to these cases due to mixed pathologies.

### ICD codes

In 429 hospitalized patients with clinically relevant RSV infection we examined the ICD-10 codes. RSV specific codes (J20.5 and J21) were only used in 4.4% of these patients. The most frequently used ICD codes were J18 (Pneumonia organism unspecified, 84 times, 19.6%) and J40-47 (Chronic lower respiratory diseases, 83 times 19.3%). ICD-codes that represented disease of the respiratory system (J00-J99) were used for 71.6% of these patients followed by 9.1% with R00-R99 codes (symptoms of the respiratory or circulatory tract) and 7.9% with I00-I99 codes (Diseases of the circulatory system). An overview of these ICD codes can be found in Table S8.

## DISCUSSION

In this study, we investigated the population risk of RSV-hospitalization and characterized patients that presented in hospital with proven RSV infection. We found that the risk of RSV-hospitalization during the RSV season was 0.04-0.24% in adults aged ≥60 years, which translates to one in 416-2500 community-dwelling older adults. Underlying comorbidities increase this risk substantially as shown by an estimated population risk of RSV-hospitalization of 0.17-1.76% in COPD (one in 57-588 patients) and 0.14-5.0% in patients with CHD (one in 20-714 patients). This increased risk was also observed in younger adults. The prevalence of (these) comorbidities was high in patients hospitalized with RSV as compared to the population prevalence. This was also true for hospitalized adults aged 20-59 years old in which 76% had underlying comorbidities. The increased risk of severe disease in the younger adults with comorbidity was also shown in previous literature by Osei-Yeboah and colleagues [18]. The study by Widmer and colleagues in 2014 used a similar approach as used in our study and found a population risk of RSV-hospitalization of 0.11% for adults aged ≥50 years although patients had to consent for participating in the study during their ED visit which could underestimate the incidence [19]. The recent study by Branche and colleagues also estimated incidence based on hospital confirmed cases and found a population risk of RSV-hospitalization of 0.14-0.26% in adults aged ≥65 years [20]. They also identified underlying cardiopulmonary disease as a significant driver of RSV-hospitalization [20]. In our current study, RSV was judged to be a clinically relevant finding in 88% of in-hospital visits. Exacerbation of underlying diseases by RSV was found to be the main reason for hospital admission. Of interest, still 25.8% of hospitalizations were judged to be caused by isolated RSV-infection. ICU admittance rate among patients hospitalized with clinically relevant RSV was 11.2% and in-hospital mortality was 8.1% although mortality was almost exclusively seen in patients aged ≥60 years. Our estimates for in-hospital severity and mortality, and presence of underlying illnesses are in line with previous literature [21, 22].

The strength of this study is the unbiased RSV testing that was routinely performed since the outbreak of COVID-19. RSV testing was never routine practice before and case definitions were used to determine who should be tested for RSV. We show that even in the hospital setting, fever is not a valid symptom to trigger respiratory testing for RSV as only 38% of cases reported fever and only 29.9% had confirmed fever in the ED thereby fulfilling the WHO case definition of ILI/SARI [23]. This is in line with our previous study in community-dwelling older adults [12]. Another strength is the data collection from three different levels of inpatient care ensuring a representative case mix of patients presenting at the hospital with RSV. Local differences in patient characteristics between hospitals were observed which emphasizes the necessity of this multicenter approach. All hospital records were manually analyzed which made it possible to determine the role of RSV in the clinical course of disease. Often RSV plays a role in the exacerbation of underlying disease but is not captured in ICD codes which limits the sensitivity of registry studies [11]. We confirmed this by observing that only 4.4% of hospitalized patients had an RSV specific ICD code, and only 71.6% had a respiratory disease ICD-code. Another strength is the inclusion of adults aged 20-59 years old, a group in which the burden of RSV is poorly characterized but in which we show that RSV is relevant in those with underlying disease. Finally, inclusion of multiple RSV seasons, as performed in this study, is vital since differences in epidemiology vary from year-to-year.

This study has also limitations. First, even though we identified 709 inpatients with RSV, numbers were still relatively small hampering risk estimation, especially in younger adults and in those with comorbidities. Second, the incidence estimates in the Jeroen Bosch Hospital were structurally higher compared to the Flevo Hospital. Demographic age differences on a population level exist but are unlikely to have caused a difference in the fraction of disease. We cannot exclude local differences in RSV epidemiology, prevalence of comorbidity, testing procedures and/or spillover of (severe) cases to other hospitals. Third, the population prevalence estimates for comorbidity are based on national data collected from a network of general practitioners offices [15]. Although distributed throughout the Netherlands, regional differences may exist. Additionally, prevalence rates of comorbidity differ significantly per age group requiring more detailed stratification in even larger cohorts than used in this study. Fourth, our study period included two clear peaks in RSV incidence but also a smaller, extended summer peak in 2022 (Figure 1). These cases were included in the in-hospital analyses but were excluded for the evaluation of the incidence of RSV-hospitalization. We believe this is justified because it is not part of the normal epidemiology of RSV to have a summer peak (in the northern hemisphere).

RSV vaccines were effective in large phase-III trials in older adults and show to provide protection spanning multiple seasons [4-6, 24, 25]. These vaccines are FDA and EMA approved and are currently entering the market. Based on the population risk for various age groups combined with the characteristics of those that were hospitalized with RSV in our study, it seems that underlying comorbidity rather than just age should play an important role in decision making who should be eligible to receive vaccination.

## Data Availability

All data produced in the present study are available upon reasonable request to the authors

## Authors’ contributions

KK, PW and EK conceptualized and designed the study, KK, MW, TL, AW, PH contributed to the acquisition of patients, KK analysed the study results and drafted the manuscript. MW, LT, AW, PH, PW, EK critically reviewed the manuscript and approved the version to be published.

