## Supplemental data for "Unveiling the spectrum of Respiratory Syncytial Virus disease in Adults: from Community to Hospital"

### Content

**Table S1.** Role of RSV (page 2)

**Table S2**. RSV epidemiology in adults aged 20-60 years old (page 3)

**Table S3**. RSV epidemiology in adults aged 20-60 years old with comorbidity (page 4)

**Table S4.** RSV epidemiology stratified by age groups (page 5)

**Table S5.** Cohort characteristics stratified by site (page 6)

**Table S6.** Severity of infection in those with clinically relevant respiratory syncytial virus infection (RSV) stratified by age group (page 7)

**Table S7.** Characteristics of patients treated with antibiotics for presumed bacterial superinfection stratified by bacterial superinfection score (page 8)

**Table S8.** ICD codes in patients admitted with clinically relevant RSV infection (page 9)

**Table S1.** Role of RSV

| Role | Comorbidity | Radiographic imaging | Microbiology | Antibiotics | Clinic | Lab | Clinical diagnosis |
|---|---|---|---|---|---|---|---|
| **Bacterial superinfection** | All is possible | Lobar infiltrate | Sputum/BAL/blood culture with bacterial pathogen | Started and continued | Fever, cough | High CRP (>100 mg/L) | Bacterial pneumonia |
| **Exacerbation respiratory disease** | COPD/Asthma | No lobar infiltrate | No culture / culture negative | Not started, discontinued | Improvement with corticosteroid therapy | CRP <100 mg/L WBC <11 x$10^9$/L | Exacerbation COPD/Asthma |
| **Exacerbation cardiac disease** | Congestive heart failure | Pulmonary edema | No culture / culture negative | Not started, discontinued | Improvement with diuretics | CRP<100 mg/L WBC <11 x$10^9$/L NT-proBNP >300 | Decompensatio cordis (acute congestive heart failure) |
| **Primary RSV** | All is possible, immunocompromised | No lobar pneumonia | No culture / culture negative | Not started, discontinued | - | CRP<100 mg/L WBC <11 x$10^9$/L | RSV only |
| **RSV not of importance** | All is possible | - | RSV CT >32 | - | Other diagnosis and no (or little) respiratory symptoms | - | Other diagnosis |

This table is used as a guide to aid in determining the role of RSV when it is not clear from the hospital record which diagnosis is made. CRP = c-reactive protein. WBC = white bloodcell count.

**Table S2**. Respiratory syncytial virus (RSV) epidemiology in adults aged 20-60 years old

| | | **Region Flevo hospital** | | **Region Jeroen Bosch hospital** | |
|---|---|---|---|---|---|
| | | **2022-2023** | **2023-2024** | **2022-2023** | **2023-2024** |
| **Population level** | Number of people | 121778 | 124179 | 84986 | 85997 |
| | Estimated RSV incidence (n)[1] | 8524 | 8693 | 5949 | 6020 |
| **Emergency department** | RSV cases (n) | 15 | 11 | 26 | 13 |
| | Incidence from population[2] | 0.01% | 0.009% | 0.03% | 0.02% |
| | Incidence from RSV positive (%)[3] | 0.18% | 0.13% | 0.44% | 0.22% |
| **Hospitalized** | RSV cases (n) | 14 | 7 | 17 | 8 |
| | Incidence from population[2] | 0.01% | 0.006% | 0.02% | 0.009% |
| | incidence from RSV positive (%)[3] | 0.16% | 0.08% | 0.29% | 0.13% |

[1] Estimated based on a 7% RSV incidence for healthy adults aged 18-60 years (11) * the susceptible population.
[2] Number of RSV cases visiting the ED/hospitalized divided by the total number of people in this population *100%
[3] Number of RSV cases visiting the ED/ hospitalized divided by the number of estimated RSV cases in the population *100%

**Table S3**. Respiratory syncytial virus (RSV) epidemiology in adults aged 20-60 years old with comorbidity

| | | Region Flevo hospital | | Region Jeroen Bosch hospital | |
|---|---|---|---|---|---|
| | | **2022-2023** | **2023-2024** | **2022-2023** | **2023-2024** |
| **Population level** | Total number of people[1] | 121778 | 124179 | 84986 | 85997 |
| | Patients with COPD (0.3-3.6%)[2] | 365-4384 | 373-4470 | 255-3059 | 258-3096 |
| | Patients with asthma (8.6-11.5%)[2] | 10473-14004 | 10679-14281 | 7309-9773 | 7396-9890 |
| | Patients with CHD (0-0.7%%)[2] | 0-852 | 0-869 | 0-595 | 0-602 |
| | Patients with diabetes (0.6-8.7%)[2] | 731-10595 | 745-10804 | 510-7394 | 516-7482 |
| **Emergency department RSV cases** | COPD: n(incidence[3]) | 4 (0.09-1.09%) | 2 (0.04-0.53%) | 5 (0.16-1.96%) | 1 (0.03-0.39%) |
| | Asthma: n(incidence[3]) | 6 (0.04-0.06%) | 2 (0.01-0.02%) | 1 (0.01-0.014%) | 3 (0.03-0.04%) |
| | CHD: n(incidence[3]) | 0 (0%) | 2 (0-0.23%) | 1 (0-0.17%) | 1 (0-0.17%) |
| | Diabetes: n(incidence[3]) | 2 (0.02-0.27%) | 5 (0.05-0.67%) | 2 (0.03-0.39%) | 0 (0%) |
| **Hospitalized RSV cases** | COPD: n(incidence[3]) | 4 (0.09-1.09%) | 1 (0.02-0.27%) | 4 (0.13-1.57%) | 1 (0.03-0.39%) |
| | Asthma: n(incidence[3]) | 6 (0.04-0.06%) | 1 (0.005-0.01%) | 1 (0.01-0.014%) | 3 (0.03-0.04%) |
| | CHD: n(incidence[3]) | 0 (0%) | 1 (0-0.12%) | 1 (0-0.17%) | 1 (0-0.17%) |
| | Diabetes: n(incidence[3]) | 2 (0.02-0.27%) | 3 (0.03-0.40%) | 2 (0.03-0.39%) | 0 (0%) |

COPD = chronic obstructive pulmonary disease. CHD = congestive heart disease.

[1] Based on regional data. (1) [2] Prevalence based on the number of persons with that comorbidity per 1000 persons in the population, lower and upper limits define the range in prevalence within age groups between 20-60 years old (2) [3] Confirmed RSV cases from this study visiting the ED/hospital with that comorbidity divided by the total number of people in this population *100%

**Table S4.** Respiratory syncytial virus (RSV) epidemiology stratified by age groups

| Age group | Region Flevo hospital | | | | | | Region Jeroen Bosch hospital | | | | | |
|---|---|---|---|---|---|---|---|---|---|---|---|---|
| | 2022-2023 | | | 2023-2024 | | | 2022-2023 | | | 2023-2024 | | |
| | Population number[1] | RSV cases | Hospitalization risk (%) | Population number[1] | RSV cases | Hospitalization risk (%) | Population number[1] | RSV cases | Hospitalization risk (%) | Population number[1] | RSV cases | Hospitalization risk (%) |
| 20-40 | 60862 | 2 | 0.003 | 63291 | 1 | 0.002 | 42829 | 2 | 0.005 | 44036 | 1 | 0.002 |
| 40-50 | 28891 | 3 | 0.010 | 29232 | 0 | 0.000 | 19422 | 1 | 0.005 | 19252 | 2 | 0.010 |
| 50-60 | 32025 | 9 | 0.028 | 31656 | 6 | 0.019 | 22735 | 14 | 0.062 | 22709 | 5 | 0.022 |
| 60-70 | 25303 | 19 | 0.075 | 26240 | 8 | 0.030 | 19219 | 26 | 0.135 | 19500 | 17 | 0.087 |
| 70-80 | 12310 | 16 | 0.130 | 13154 | 8 | 0.061 | 13906 | 35 | 0.252 | 14493 | 17 | 0.117 |
| 80-90 | 3857 | 7 | 0.181 | 4017 | 3 | 0.075 | 5667 | 26 | 0.459 | 5775 | 18 | 0.312 |
| >90 | 696 | 5 | 0.718 | 696 | 0 | 0.000 | 1015 | 7 | 0.690 | 992 | 5 | 0.504 |

[1] Based on regional data (1)

**Table S5.** Characteristics of the study cohort stratified by site

| | **Amsterdam University Medical Center N=296** | **Jeroen Bosch Hospital N=291** | **Flevo Hospital N=122** |
|---|---|---|---|
| Male | 149 (50.3%) | 130 (44.7%) | 52 (42.6%) |
| Age (median, [IQR])<br>- 20-39<br>- 40-59<br>- 60-79<br>- 80 and older | 67 [51-75]<br>- 41 (13.9%)<br>- 66 (22.4%)<br>- 136 (46.1%)<br>- 52 (17.6%) | 74 [64-82]<br>- 9 (3.1%)<br>35 (12.0%)<br>156 (53.6%)<br>91 (31.3%) | 68 [59-76]<br>- 6 (5.0%)<br>- 24 (19.8%)<br>- 66 (54.5%)<br>- 25 (20.7%) |
| Any comorbidity[1] | 253 (85.5%) | 240 (82.5%) | 100 (82.0%) |
| Pulmonary disease<br>- COPD<br>- Asthma | 111 (37.5%)<br>- 55 (18.6%)<br>- 39 (13.2%) | 142 (48.8%)<br>- 99 (34.0%)<br>- 31 (10.7%) | 68 (55.7%)<br>- 45 (36.9%)<br>- 20 (16.4%) |
| Congestive heart disease (CHD) | 57 (19.3%) | 67 (23.0%) | 17 (13.9%) |
| Active malignancy<br>- Hematologic<br>- Solid tumor | 73 (24.7%)<br>- 44 (14.9%)<br>- 30 (10.1%) | 43 (14.8%)<br>- 17 (5.8%)<br>- 25 (8.6%) | 11 (9.0%)<br>- 5 (4.1%)<br>- 6 (4.9%) |
| Metabolic disease<br>- Diabetes<br>- Renal insufficiency (hemodialysis) | 94 (31.8%)<br>- 54 (18.2%)<br>- 8 (2.7%) | 84 (28.9 %)<br>- 62 (21.3%)<br>- 4 (1.4%) | 39 (32.0%)<br>- 28 (23.0%)<br>- 3 (2.5%) |
| Organ transplant[2] | 10 (3.4%) | 4 (1.4%) | 1 (0.8%) |
| Rheumatic / immunologic disease[2] | 45 (15.2%) | 36 (12.4%) | 18 (14.8%) |
| Hospitalization | 189 (63.9%) | 214 (73.5%) | 100 (82.0%) |
| Role of RSV<br>- Primary RSV infection<br>- Factor in exacerbation of underlying disease<br>- Bacterial superinfection after RSV<br>- RSV was not of importance<br>- Indeterminate[3] | <br>- 121 (40.9%)<br>- 93 (31.4%)<br>- 37 (12.5%)<br>- 44 (14.9%)<br>- 1 (0.3%) | <br>- 89 (30.6%)<br>- 108 (37.1%)<br>- 61 (21.0%)<br>- 31 (10.7%)<br>- 2 (0.6%) | <br>- 32 (26.2%)<br>- 49 (40.2%)<br>- 27 (22.2%)<br>- 12 (9.8%)<br>- 2 (1.6) |

[1.] From the comorbidities that were noted in the method section [2.] With active immunosuppressive therapy or illness. [3] No definitive role of RSV could be addressed to these cases due to mixed pathologies.

**Table S6.** Severity of infection in those with clinically relevant respiratory syncytial virus infection (RSV) stratified by age group

| | **20-59 years**<br>**n=76** | **60 years and older**<br>**n=358** |
|---|---|---|
| Length of stay median days [IQR] | 4 [2-7] | 5 [3-8] |
| Treated with antibiotics[1] | 45 (59.2%) | 248 (69.9%) |
| Infiltrate found with chest radiography | 23 (30.3%) | 141 (39.5%) |
| Bacterial culture positive | 17 (37.8%) | 45 (18.1%) |
| Oxygen therapy n(%)<br>median days [IQR] | 50 (67.6%)<br>4 [2-7] | 281 (78.5%)<br>3 [2-7] |
| Intensive care unit admission n(%)<br>median days in the ICU [IQR] | 14 (18.9%)<br>12 [5-19] | 34 (9.6%)<br>3 [2-8] |
| Invasive ventilation n(%)<br>median days [IQR] | 9 (12.0%)<br>9 [8-11] | 15 (4.2%)<br>4 [3-7] |
| In-hospital mortality | 2 (2.7%) | 33 (9.3%) |

IQR = interquartile range**.** [1.] For presumed respiratory bacterial superinfection

**Table S7.** Characteristics of patients treated with antibiotics for presumed bacterial superinfection stratified by bacterial superinfection score

| | **Unlikely (score <2) n=108** | **Possible (score 2-3) n=44** | **Likely (score >3) N=141** |
|---|---|---|---|
| Age (median, [IQR])<br>- 20-39<br>- 40-59<br>- 60-79<br>- 80 and older | 72 [66-82]<br>- 3 (2,8%)<br>- 12 (11,1%)<br>- 57 (52,8%)<br>- 36 (33,3%) | 69 [60-74]<br>- 3 (6,8%)<br>- 7 (15,9%)<br>- 28 (63,6%)<br>- 6 (13,6%) | 72 [66-82]<br>- 3 (2,1%)<br>- 17 (12,1%)<br>- 74 (52,5%)<br>- 47 (33,3%) |
| Any comorbidity[1] | 100 (92,6%) | 39 (88,6%) | 115 (81,6%) |
| Pulmonary disease<br>- COPD<br>- Asthma | 64 (59,3%)<br>- 41 (38,0%)<br>- 12 (11,1%) | 22 (50,0%)<br>- 15 (34,1%)<br>- 5 (11,4%) | 64 (45,4%)<br>- 44 (31,2%)<br>- 17 (12,1%) |
| Congestive heart failure | 25 (23,1%) | 11 (25,0%) | 30 (21,3%) |
| Active malignancy<br>- Hematologic<br>- Solid tumor | 17 (15,7%)<br>- 8 (7,4%)<br>- 8 (7,4%) | 5 (11,4%)<br>- 3 (6,8%)<br>- 2 (4,5%) | 27 (19,1%)<br>- 16 (11,3%)<br>- 11 (7,8%) |
| Metabolic disease<br>- Diabetes<br>- Renal insufficiency (hemodialysis) | 44 (40,7%)<br>- 33 (30,6%)<br>- 3 (2,8%) | 16 (36,4%)<br>- 14 (31,8%)<br>- 0 (0,0%) | 35 (24,8%)<br>- 23 (16,3%)<br>- 2 (1,4%) |
| Organ transplant[2] | 1 (0,9%) | 1 (2,3%) | 3 (2,1%) |
| Rheumatic / immunologic disease[2] | 14 (13,0%) | 5 (11,4%) | 19 (13,5%) |
| **Vital signs and laboratory studies in the ED** | | | |
| Oxygen saturation ED (median [IQR]) | 90 [87-95] | 93 [88-95] | 90 [86- 95] |
| Respiratory rate (bpm, median [IQR]) | 24 [20-28] | 25 [20-28] | 22 [19- 28] |
| Pulse (bpm, median [IQR]%) | 98 [85-110] | 105 [85-121] | 101 [85-115] |
| Temperature (°C, median [IQR])<br>Fever measured in the ED (≥38°C) | 37,5 [36,8- 38,3]<br>36 (34,6%) | 37,7 [37,0- 38,6]<br>19 (43,2%) | 37,8 [36,9-38,5]<br>65 (47,1%) |
| CRP mg/L (median [IQR]) | 66 [32-112] | 36 [25-56] | 135 [48-236] |
| Leukocyte count x10^9/L (median [IQR]) | 9,6 [6,9-12,6] | 9,1 [7,1-10,4] | 13,3 [9,8-15,9] |
| **Hospitalization severity** | | | |
| Infiltrate found on chest radiography | 0 (0,0%) | 36 (81,8%) | 125 (88,7%) |
| Bacterial culture positive | 0 (0,0%) | 8 (18,2%) | 54 (38,3%) |
| Oxygen therapy | 82 (75,9%) | 34 (77,3%) | 117 (83,0%) |
| Intensive care admission | 12 (11,1%) | 5 (11,4%) | 21 (14,9%) |
| Invasive ventilation | 3 (2,8%) | 3 (6,8%) | 12 (8,6%) |
| Mortality | 9 (8,3%) | 5 (11,4%) | 12 (8,5%) |
| Role of RSV<br>- Primary RSV infection<br>- Factor in exacerbation of underlying disease<br>- Bacterial superinfection after RSV<br>- Indeterminate[3] | <br>49 (45,4%)<br>55 (50,9%)<br>3 (2,8%)<br>1 (0,9%) | <br>9 (20,5%)<br>16 (36,4%)<br>18 (40,9%)<br>1 (2,3%) | <br>12 (8,5%)<br>33 (23,4%)<br>95 (67,4%)<br>1 (0,7%) |

IQR = interquartile range [1.] From the comorbidities that were noted in the method section [2.] With active immunosuppressive therapy or illness. [3] No definitive role of RSV could be addressed to these cases due to mixed pathologies.

**Table S8.** ICD codes in patients admitted with clinically relevant RSV infection

| ICD-code | Primary RSV infection N=112 | Factor in exacerbation of underlying disease N=201 | Bacterial super-infection after RSV N=116 | Total N=429 |
|---|---|---|---|---|
| J00-J99 Diseases of respiratory system | 71 (63.4%) | 149 (74.1%) | 87 (75.0%) | 307 (71.6%) |
| - J00-06 Acute Upper RTI | - 1 (0.9%) | - 0 (0%) | - 0 (0%) | - 1 (0.2%) |
| - J09-12 Viral pneumonia | - 11 (9.8%) | - 15 (7.5%) | - 11 (9.5%) | - 37 (8.6%) |
| - J13-15 Bacterial pneumonia | - 1 (0.9%) | - 1 (0.5%) | - 9 (7.8%) | - 11 (2.6%) |
| - J18 Pneumoniae organism unspecified | - 22 (19.6%) | - 10 (5.0%) | - 52 (44.8%) | - 84 (19.6%) |
| - J20-22 Other Acute LRTI | - 18 (16.1%) | - 40 (19.9%) | - 5 (4.3%) | -63 (14.7%) |
| - J20.5/21 RSV LRTI* | - 13 (11.6%) | - 4 (2.0%) | - 2 (1.7%) | -19 (4.4%) |
| - J40-47 Chronic lower respiratory diseases | - 1 (0.9%) | - 77 (38.3%) | - 5 (4.3%) | -83 (19.3%) |
| - J80-99 Other respiratory diseases** | - 3 (2.7%) | - 5 (2.5%) | - 3 (2.7%) | -11 (2.6%) |
| I00-I99 Diseases of circulatory system | 3 (2.7%) | 28 (13.9%) | 3 (2.6%) | 34 (7.9%) |
| - I20-25 Ischemic heart diseases | - 0 (0%) | - 0 (0%) | - 2 (1.7%) | - 2 (0.5%) |
| - I30-52 Other forms of heart disease | - 2 (1.8%) | - 24 (11.9%) | - 1 (0.9%) | - 27 (6.3%) |
| - Other I00-I99*** | - 1 (0.9%) | - 4 (2%) | - 0 (0%) | - 5 (1.2%) |
| R00-R99 Symptoms, signs and abnormal clinical and laboratory findings, not elsewhere classified | 21 (18.7%) | 9 (4.5%) | 9 (7.7%) | 39 (9.1%) |
| - R00-09 Involving circulatory and respiratory | - 8 (7.1%) | - 7 (3.5%) | - 5 (4.3%) | - 20 (4.7%) |
| - R50-69 General symptoms and signs | - 13 (11.6%) | - 2 (1.0%) | - 4 (3.4%) | - 19 (4.4%) |
| A00-B99 Infectious and parasitic diseases | 0 (0%) | 0 (0%) | 3 (2.6%) | 3 (0.7%) |
| - A30-49 Other bacterial diseases | 0 (0%) | 0 (0%) | 3 (2.6%) | - 3 (0.7%) |
| C00-D48 Neoplasms | 4 (3.6%) | 0 (0%) | 1 (0.9%) | 5 (1.2%) |
| - C00-97 Malignant neoplasms | 4 (3.6%) | 0 (0%) | 1 (0.9%) | - 5 (1.2%) |
| D50-D89 Diseases of the blood and blood-forming organs and certain disorders involving the immune mechanism | 5 (4.5%) | 2 (1.0%) | 1 (0.9%) | 8 (1.9%) |
| E00-E90 Endocrine, nutritional and metabolic diseases | 0 (0%) | 2 (1.0%) | 0 (0%) | 2 (0.5%) |
| G00-G99 Diseases of the nervous system | 1 (0.9%) | 0 (0%) | 0 (0%) | 1 (0.2%) |
| N00-N99 Diseases of the genitourinary system | 0 (0%) | 1 (0.5%) | 1 (0.9%) | 2 (0.5% |
| S00-T98 Injury, poisoning and certain other consequences of external causes Unknown | 1 (0.9%) | 0 (0%) | 1 (0.9%) | 2 (0.5%) |
| Z00-99 Factors influencing health status and contact with health services | 0 (0%) | 0 (0%) | 1 (0.9%) | 1 (0.2%) |
| ICD-code unknown | 15 (13.4%) | 8 (4.0%) | 10 (8.6%) | 33 (7.7%) |

* Specific code for acute bronchitis/bronchiolitis due to respiratory syncytial virus (also included in J20-22 group)
** Combination of ICD codes for other respiratory diseases of the interstitium (J80-84), suppurative and necrotic conditions (J85-86), pleural disease (J90-94), other respiratory diseases (J95-99) and COVID infection (U07.1)
*** Combination of ICD codes for other circulatory diseases including pulmonary heart disease and circulation (I26-28), Cerebrovascular diseases (I60-69) and diseases of arteries, arterioles and capillaries (I70-79).
